# Cytokine biomarkers of COVID-19

**DOI:** 10.1101/2020.05.31.20118315

**Authors:** Hai-Jun Deng, Quan-Xin Long, Bei-Zhong Liu, Ji-Hua Ren, Pu Liao, Jing-Fu Qiu, Xiao-Jun Tang, Yong Zhang, Ni Tang, Yin-Yin Xu, Zhan Mo, Juan Chen, Jie-Li Hu, Ai-Long Huang

## Abstract

We used a new strategy to screen cytokines associated with SARS-CoV-2 infection. Cytokines that can classify populations in different states of SARS-CoV-2 infection were first screened in cross-sectional serum samples from 184 subjects by 2 statistical analyses. The resultant cytokines were then analyzed for their interrelationships and fluctuating features in sequential samples from 38 COVID-19 patients. Three cytokines, M-CSF, IL-8 and SCF, which were clustered into 3 different correlation groups and had relatively small fluctuations during SARS-CoV-2 infection, were selected for the construction of a multiclass classification model. This model discriminated healthy individuals and asymptomatic and nonsevere patients with accuracy of 77.4% but was not successful in classifying severe patients. Further searching led to a single cytokine, hepatocyte growth factor (HGF), which classified severe from nonsevere COVID-19 patients with a sensitivity of 84.6% and a specificity of 97.9% under a cutoff value of 1128 pg/ml. The level of this cytokine did not increase in nonsevere patients but was significantly elevated in severe patients. Considering its potent antiinflammatory function, we suggest that HGF might be a new candidate therapy for critical COVID-19. In addition, our new strategy provides not only a rational and effective way to focus on certain cytokine biomarkers for infectious diseases but also a new opportunity to probe the modulation of cytokines in the immune response.

## Introduction

The pandemic of coronavirus disease 2019 (COVID-19) is imposing a heavy burden on global public health. As of May 30, 2020, COVID-19 has been confirmed in more than 5 million people worldwide, carrying a mortality of approximately 6.2% [1]. COVID-19 is caused by infection with severe acute respiratory syndrome coronavirus-2 (SARS-CoV-2), which shares 79.5% sequence homology with SARS-CoV [2]. The high transmission capability of SARS-CoV-2, which is indicated by a basic reproductive number R_0_ of 2.0 – 3.77 [3, 4, 5], led to the rapid spread of the virus from a city to the whole world in 3 months.

SARS-CoV-2 uses angiotensin-converting enzyme 2 (ACE2) as a receptor to infect cells [6, 7]. The ACE2 distribution is enriched in human alveolar epithelial cells, making the lung a vulnerable target organ [8]. Patients infected with SARS-CoV-2 commonly manifest lower respiratory tract symptoms of dry cough or, less commonly, dyspnea [9, 10]. These symptoms reflect, to some extent, inflammation in the lung. Inflammation is a ‘side effect’ of the immune response. Viral infection induces innate and/or adaptive immune responses by the host immune system. Initiation of these responses can result in the production of some cytokines that induce a proinflammatory response and attract cells, such as neutrophils and macrophages, to sites of infection and in turn cause damage to normal host tissues by releasing cytotoxic substances [11]. In COVID-19 patients, evidence has indicated that the levels of some cytokines are significantly elevated in COVID-19 patients, and cytokine storms are associated with the severity of the disease [10, 12, 13, 14]. In this study, we searched cytokine biomarkers of COVID-19 by a new strategy taking into account the fluctuations and interrelationships of cytokines, which have been largely overlooked in previous studies, and identified cytokines that can classify populations with different conditions during SARS-CoV-2 infection.

## Methods

### Patients and samples

Cross-sectional serum samples were obtained from 184 subjects, including 37 healthy controls, 37 asymptomatic individuals infected with SARS-CoV-2 confirmed by RT-PCR, 97 nonsevere or moderate COVID-19 inpatients, and 13 severe COVID-19 patients. The asymptomatic individuals and COVID-19 inpatients were included from previous studies. Asymptomatic individuals and COVID-19 patients were confirmed by RT-PCR assay from nasal and pharyngeal swab specimens.

Sequential serum samples used for the fluctuation analysis were obtained from 38 COVID-19 inpatients. These patients were among the subjects in a previous study [15]. Serum samples were collected from these patients every 3 days during hospitalization, and each patient provided at least 4 sequential samples.

The study was approved by the Ethics Commission of Chongqing Medical University (reference number: 2020006–1). Written informed consent was waived by the Ethics Commission of the designated hospital for emerging infectious diseases.

### Detection of cytokines

Concentrations of 48 cytokines and chemokines in serum samples were measured using a Bio-Plex Pro Human Cytokine Screening Panel (48-Plex #12007283, Bio-Rad) according to the manufacturer’s instructions. All serum samples were inactivated at 56 °C for 30 min before the assay.

### Definitions

A confirmed case of SARS-CoV-2 infection was defined as an individual with nasopharyngeal swabs positive for SARS-CoV-2 nucleic acid by RT-PCR. An asymptomatic case was defined as an individual with a positive RT-PCR result but without any relevant clinical symptoms in the preceding 14 days but before 2 consecutive negative RT-PCR results. Severe COVID-19 was determined if a COVID-19 patient showed any of the following: shortness of breath, RR ≥ 30 bpm, blood oxygen saturation ≤ 93% (at rest), PaO_2_/FiO_2_ ≤ 300 mmHg, or pulmonary inflammation that significantly progressed > 50% within 24 to 48 hours. Nonsevere patients, including mild and moderate patients, were defined as COVID-19 patients with symptoms but that could not be classified as severe.

### Multiclass classification model

A multinomial logistic regression model was used to determine the class of individuals. A multiclass classifier was trained by fitting the multinomial logistic model using the ‘*nnet*’ package of *R* software, version 3.6.0. A ten-fold cross-validation method was used to evaluate the performance of the model. Briefly, all the subjects or healthy controls (the cross-sectional samples) were randomly divided into 5 sets using the ‘*caret’* package of *R* software; 3 of the 5 sets were used for training, and the remaining 2 were used for testing. This procedure was repeated 10 times so that each set was under the testing set. The classification accuracy was assessed in 10-fold cross-validation, and the mean and standard deviation (sd) were calculated.

### Model evaluation by quasiindependent data

Serum samples collected at other timepoints from a portion of the subjects in the cross-sectional study, including 35 asymptomatic subjects, 65 nonsevere patients and 1 severe patient, were used as a quasiindependent dataset to evaluate the predictive performance of the multiclass classification model. These data were input into the model, allowing the model to predict the class of each sample in the quasiindependent data. The area under the curve (AUC) of each class was calculated to assess the performance of the model using the ‘*pROC’* package of *R* software.

### Statistical analysis

Continuous variables were expressed as the median values (interquartile ranges, IQRs), and the Mann-Whitney U test was used for comparisons; categorical variables were expressed as numbers (percentages), and the χ² test or Fisher’s exact test was used for comparisons. *p* < 0.05 was considered to indicate statistically significant differences, and *p* < 0.001 was considered to indicate highly statistically significant differences (shown as ***). Statistical analyses were performed using *R* software, version 3.6.0.

## Results

### Strategy for searching cytokine biomarkers for COVID-19

Ideal cytokine biomarkers for COVID-19 should meet the following 3 criteria: (1) accurate, differentiating people with different statuses of SARS-CoV-2 infection with good sensitivity and specificity; (2) stable, accurately differentiating these people at different stages during the infection; and (3) small in number, fewer cytokines means less expense and easier establishment of a testing system. With these criteria in mind, we used a screening strategy for searching cytokine biomarkers for COVID-19. In the 1^st^ round of screening, cross-sectional serum samples from people with different SARSCoV-2 infection statuses were tested for 48 cytokines. These cytokines were statistically tested by Wilcoxon analysis or/and ANOVA for their capacity to classify different groups. Overlapping cytokines meeting the screening criteria (described below) in the statistical analyses were selected for further screening. In the 2^nd^ round of screening, the fluctuation and similarity in the fluctuation patterns of cytokines during illness were analyzed in sequential samples. The rationale for this step is as follows. (1) A smaller fluctuation (in a patient) means more stability over time, translating to less impact of the sampling timepoint on the level of cytokines. A prerequisite for this notion is that the levels of cytokines do fluctuate, which is evidenced by the following results. (2) A similar fluctuation pattern for different cytokines indicates that these cytokines are partially redundant, at least statistically, in classifying different populations. For an extreme example, simultaneously selecting 2 cytokines with 100% correlation would be completely redundant, contradicting criterion (3) for ideal cytokine biomarkers. Cytokines selected through the 2 rounds of screening were used to construct a multiclass classification model. Then, the obtained model was tested by partially dependent samples (Fig. 1).

**Fig. 1.**
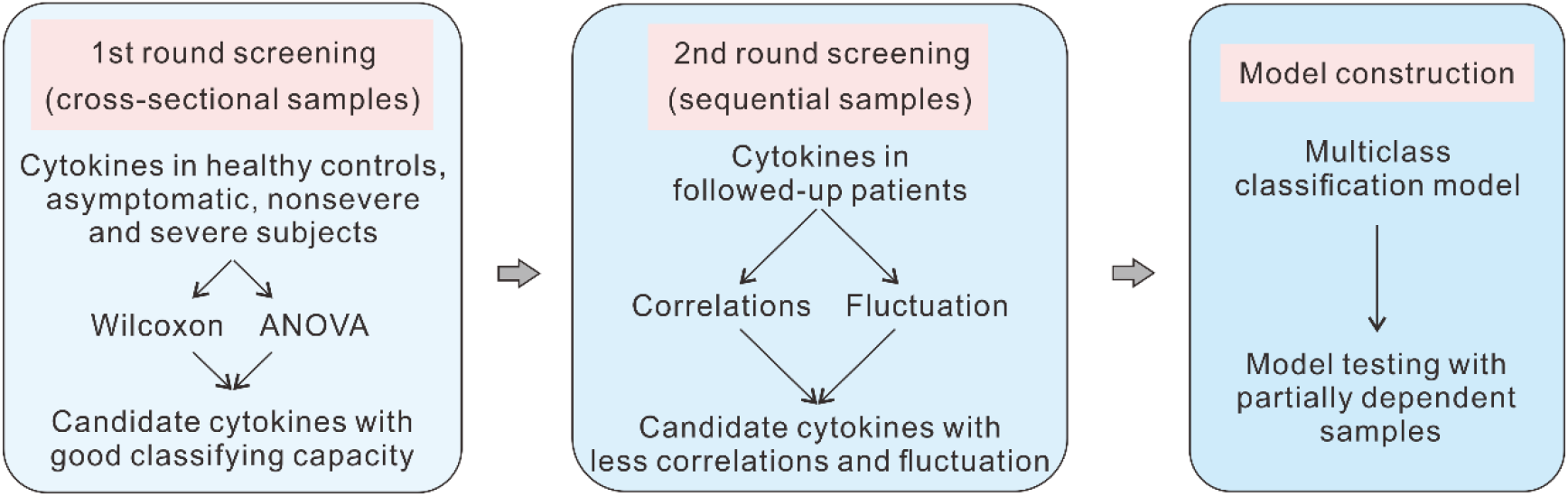
Strategy for screening cytokine biomarkers of COVID-19. The figure shows the procedure to screen cytokine biomarkers of COVID-19.

### Preliminary screening of cytokines in the cross-sectional samples

Serum samples collected from 184 subjects were measured for the concentration of 48 cytokines (Fig. 2A) by the system proven to be stable and repeatable (Fig. S1). Demographics and baseline characteristics of the subjects are summarized in Table S1. Wilcoxon analysis was used to identify cytokines with significant differences between the healthy control group and the other 3 groups. Eighteen cytokines were screened out by P<0.001 (Fig. 2B). ANOVA was used to screen cytokines that could classify the 3 groups, and 22 cytokines were selected by the criteria of P < 0.001 and F > 10 (Fig. 1C). Overlapping of the 2 sets of cytokines resulted in 10 cytokines, IL-6, IL-7, IL-8, IL-13, MIP-1α, eotaxin, M-CSF, IFN-γ, SCF and IL-R2α (Fig. 1D).

**Fig. 2.**
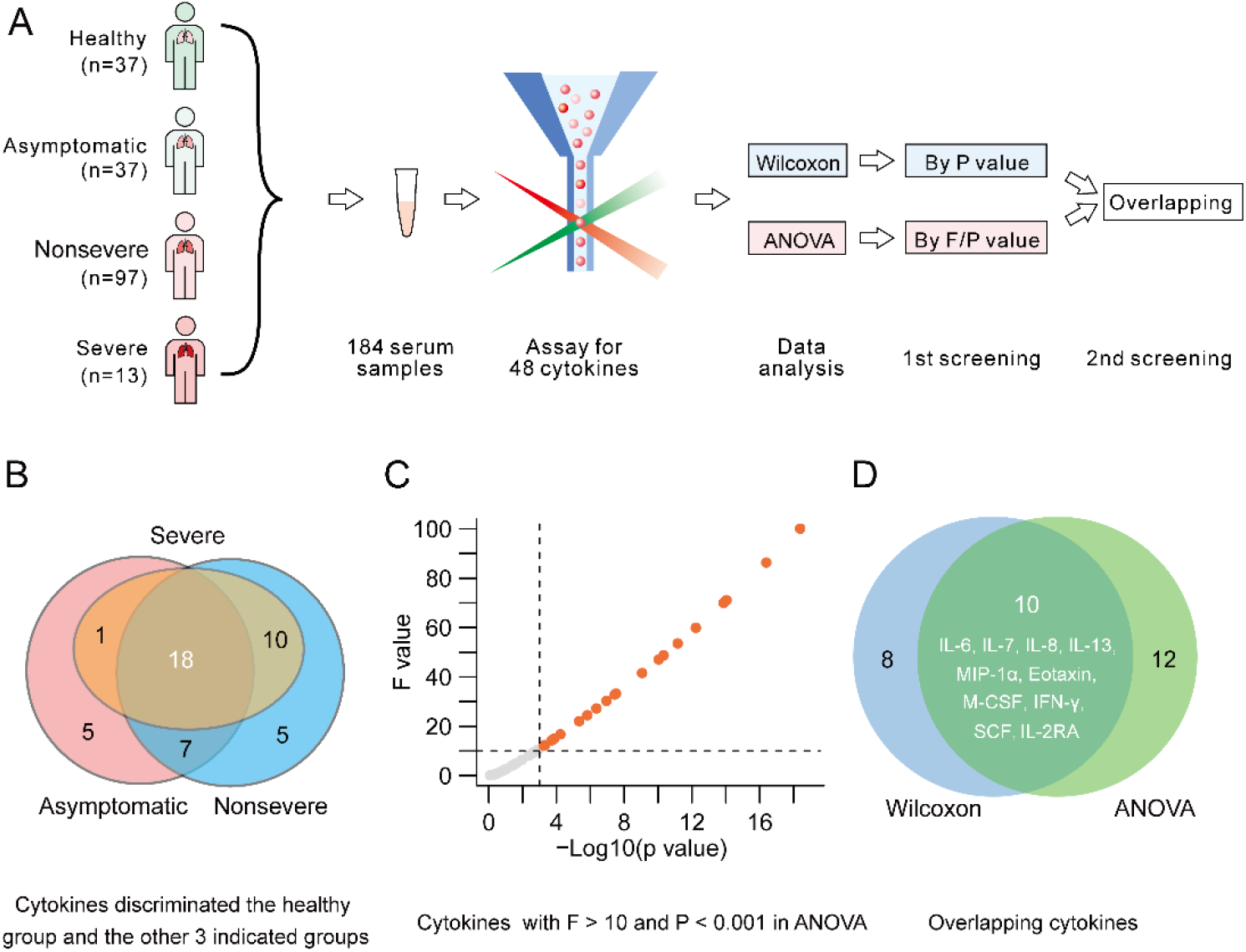
Preliminary screening of cytokines. A. Procedure of preliminary screening of candidate cytokines. B. Cytokines with the potential to discriminate the healthy control group and the other 3 groups. C. Cytokines with F > 10 and P < 0.01 in ANOVA. D. Overlapping cytokines meeting the criteria of the 2 statistical analyses.

### Screening of cytokines by using sequential samples

We followed up 38 confirmed COVID-19 patients to examine the dynamic change in 48 cytokines (Fig. 3A). The demographics and baseline characteristics of these patients are summarized in Table S2. Forty-eight cytokines in all the serum samples were detected in parallel, and the samples from one patient were detected in the same plate. The correlations among the dynamic changes in the 10 cytokines selected above were analyzed. To this end, we first normalized the concentrations of cytokines by dividing the concentrations of each cytokine at sequential points by the concentration of that cytokine at the first point (Fig. S2A). This step eliminated the variation in cytokine levels among different individuals. Correlation coefficients were calculated between each cytokine and each of the other 9 cytokines for each patient, leading to 38 correlation coefficients (38 patients) for each correlation pair. The means of these correlation coefficients were calculated and used to cluster the cytokines. As shown in Fig. 3B, 3 clusters of correlated cytokines were found. Cluster 1 includes IL-6 and M-CSF. Cluster 2 includes IL-7 and IL-8. Cluster 3 comprises IL-13, SCF, MIP-1α, eotaxin, IFN-γ and IL-2Rα. Relatively high correlations were found for the cytokine pairs IL-6/M-CSF (r = 0.79) and IFN-γ/IL-2Rα (r = 0.73). As examples, the fluctuating patterns of these two pairs of cytokines from 9 patients are shown in Fig. S3.

**Fig. 3.**
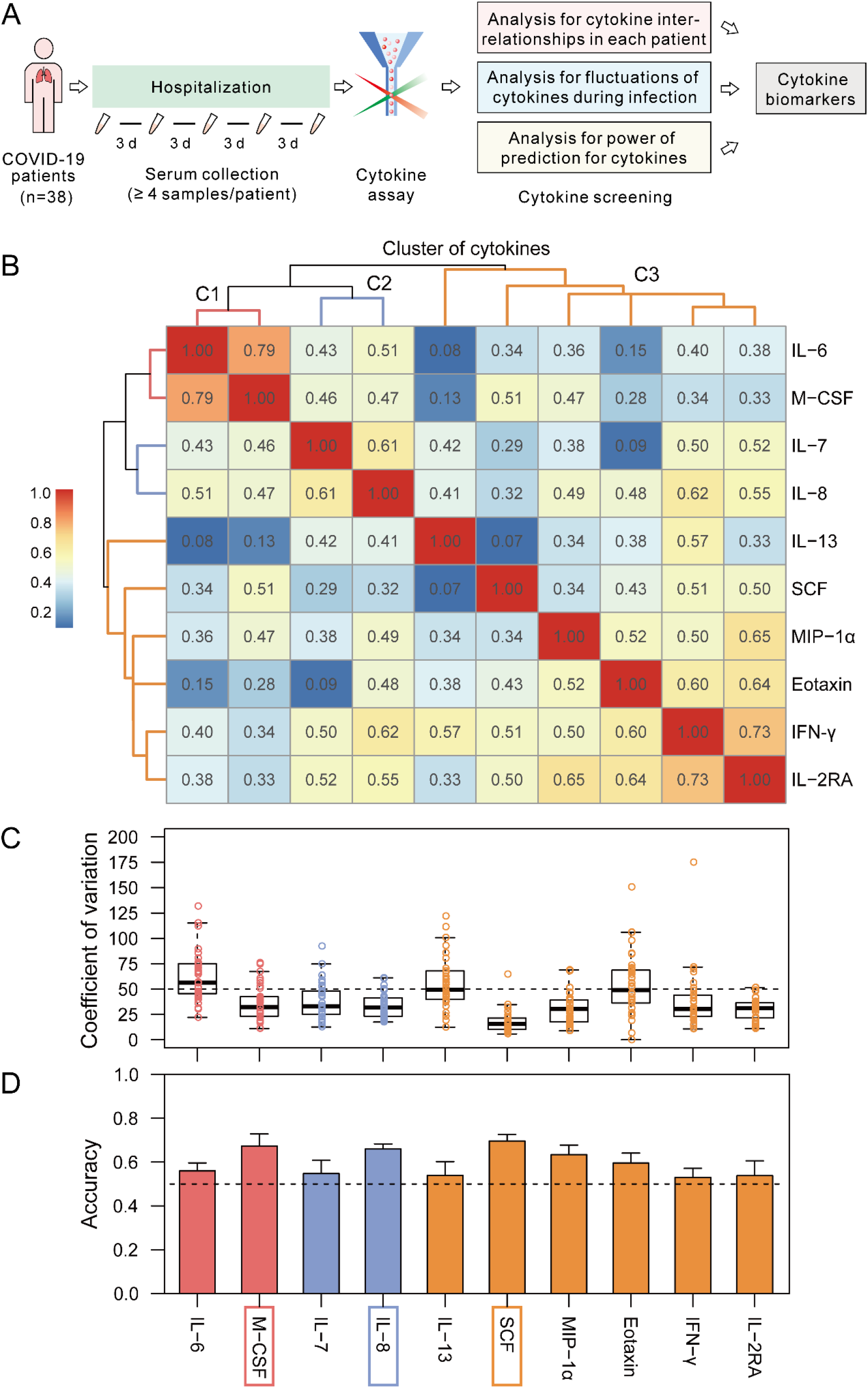
Screening of cytokines by using sequential samples. A. Procedure of cytokine screening. B. Correlation analysis of the 13 cytokines. C. CV of the levels of the 13 cytokines during hospitalization. D. Accuracy of the cytokines in classifying populations with different states of SARS-CoV-2 infection.

Choosing 1 cytokine from each cluster would decrease redundancy since the correlations within the clusters were higher than those between clusters (Fig. 3B). To target the candidate cytokines, we analyzed the coefficient of variation (CV) of the cytokines in each cluster. The CV of each cytokine for each patient was calculated using the normalized concentrations of each cytokine described above, and the CVs of each cytokine from the 38 patients are plotted in Fig. 3C. All of the cytokines fluctuated in the sequential serum samples, as reflected by median CVs ranging from 15.5% (IQR 5.8%-64.6%) (SCF) to 56.4% (IQR 22.1%-131.7%) (IL-6). The ranks of the mean CVs in each cluster are as follows: C1, IL-6 > M-CSF; C2, IL-7 > IL-8; and C3, IL-13 > eotaxin > IL-2Rα > MIP-1α > IFN-γ > SCF. The most stable cytokines in each cluster were M-CSF, IL-8 and SCF. Interestingly, these 3 cytokines also performed best in each cluster in classifying different populations by univariate multiclass classification model (Fig. 3D). Thus, we chose these cytokines for the construction of a multiclass classification model.

### Construction and testing of a multiclass classification model

The training dataset from the cross-sectional samples was used to construct a multiclass classification model. The estimated overall accuracy of classification in 10-fold cross-validation was 77.4% (with a standard deviation of 5.2%). The estimated mean accuracy for the classification of the healthy control, asymptomatic and nonsevere groups in 10-fold cross-validation was 76.0%, 76.2 and 88.6%, respectively (Fig. 4A). Unfortunately, the model performed poorly for the classification of the severe group (Fig. 4A). The AUC calculated using the quasiindependent dataset of validation was 0.883, 0.920, 0.920 and 0.923 for the healthy control, asymptomatic, nonsevere and severe groups, respectively (Fig. 4B).

**Fig. 4.**
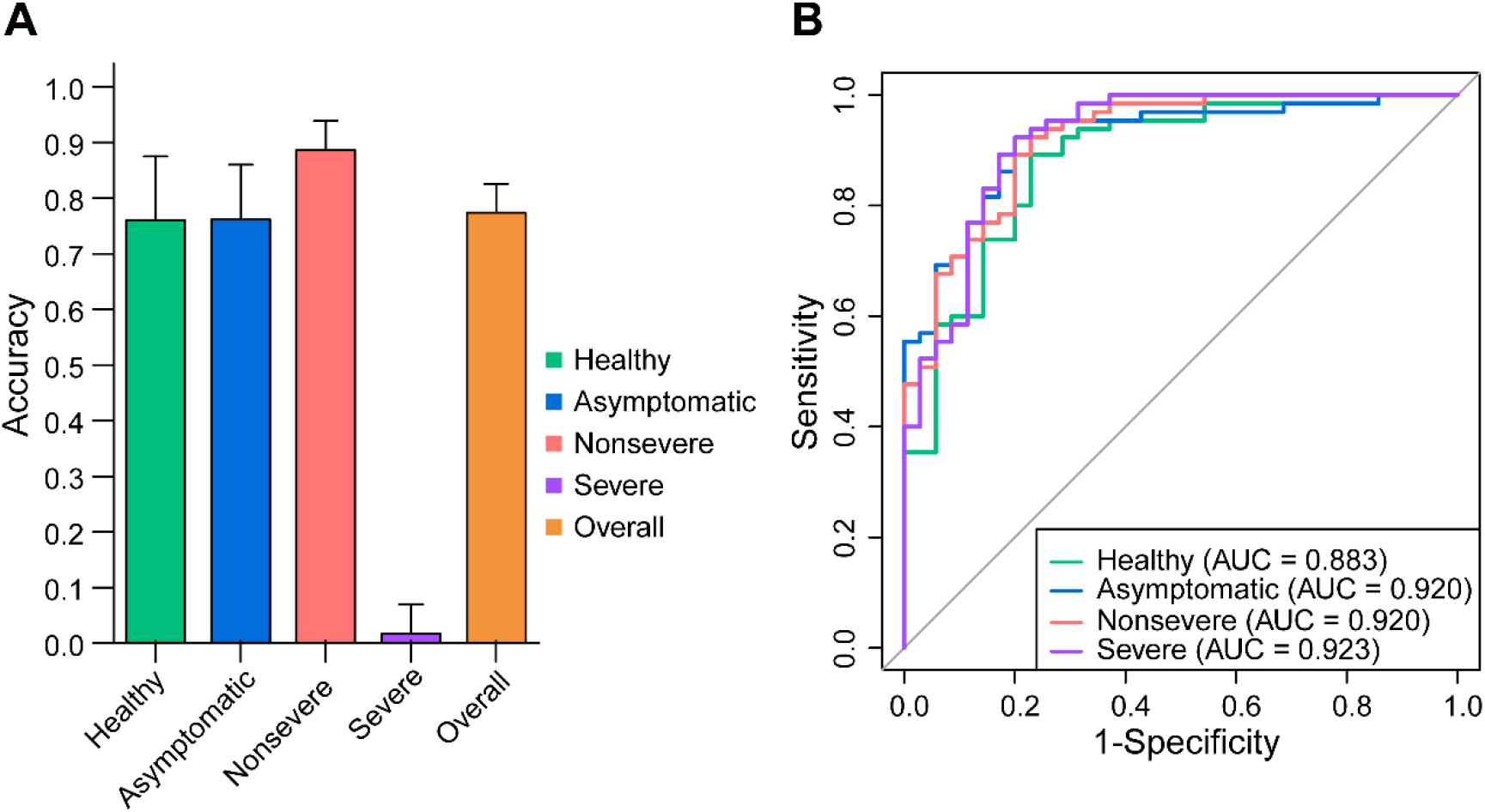
Performance of the multiclass classification model. A. Performance of the model in the training dataset. B. Performance of the model in the quasiindependent dataset.

### Identification of cytokines discriminating severe from nonsevere COVID-19

To identify cytokines with better performance in discriminating severe from nonsevere COVID-19 patients, we screened the cytokines again from among those showing significant differences between these 2 groups. Seven cytokines, IL-8, MIP-1α, HGF, IL-16, IL-18, MCP-3 and SCGF-β, were screened out by Wilcoxon analysis. Among the 7 cytokines, HGF was apparently superior to the others in ANOVA, with a far greater F value than all the other cytokines (Fig. 5B). The accuracy of classification of severe case was also the highest when using HGF (Fig. 5C). Thus, HGF was ultimately used as the biomarker to discriminate severe from nonsevere COVID-19 patients. The sensitivity and specificity of HGF to classify severe patients were 84.6% and 97.9%, respectively, with an AUC of 90.5% (Fig. 5D). The cutoff value of HGF determined by the ROC curve was 1128 pg/ml. This result means that a COVID-19 patient with an HGF concentration over 1128 pg/ml has a great possibility of being severely ill. This cutoff classified falsely only 2 out of 97 nonsevere patients and 2 out of 13 severe patients (Fig. 5E). In addition, HGF levels were not associated with age within different groups (Fig. S4) and no significant correlation was found between the fluctuation pattern of HGF and those of other cytokines (data not shown), indicating HGF is a relatively independent factor.

**Fig. 5.**
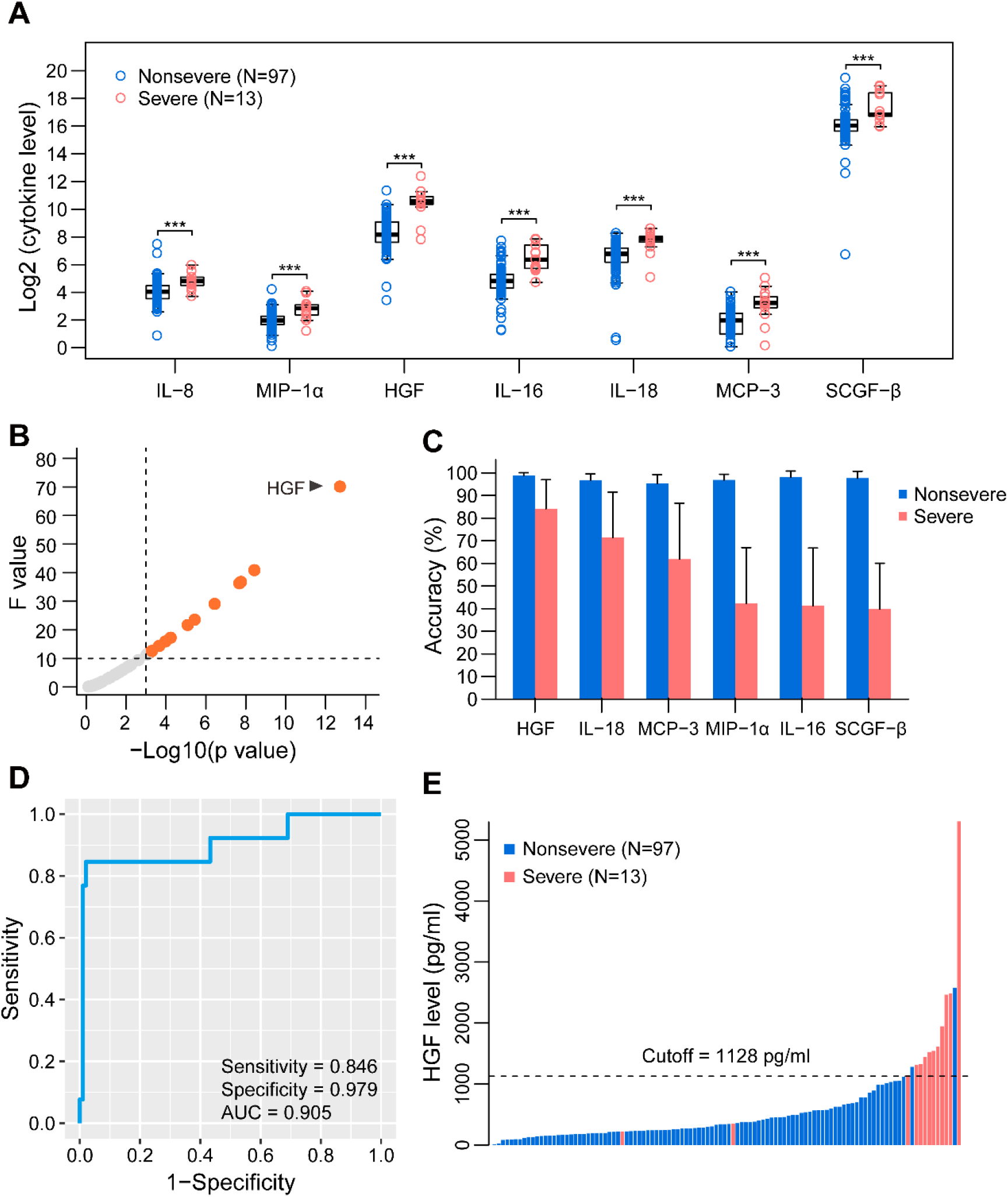
Screening of cytokines discriminating severe from nonsevere COVID-19. A, Cytokines at different concentrations in nonsevere and severe COVID-19 patients. B, Evaluation of the cytokines for their classification capacity by binomial regression model. C, Accuracy of the cytokines in classifying the 2 groups. D, ROC curve of HGF. E, Performance of the HGF cutoff value.

## Discussion

Cytokines with significant differences in populations with different states of SARS-

CoV-2 infection, including healthy controls and asymptomatic infection, nonsevere and severe COVID-19 patients, might be involved in the immune or inflammatory response induced by the virus. In our first step of searching for such cytokines, we identified 10 in cross-sectional samples by using 2 statistical analyses. To narrow down candidates, we dissected the correlations among these cytokines after noticing the fluctuation of cytokines in the followed-up COVID-19 patients.

The fluctuation in cytokine levels observed cannot be explained by a variation in the measurement. We assayed the sequential samples from a patient in one plate, and the within-run CVs (in one plate) of all the cytokine controls were less than 10% (Fig. S1), while the CV means of the 10 cytokines from the samples ranged from 15.5% to 56.4%. Indeed, cytokine fluctuation was clearly observed in a previous study, which showed that IL-6, IP-10, MCP-1 and IL-15 levels fluctuated significantly over several days in subjects experimentally infected with influenza A virus [16]. During chronic hepatitis B virus infection, some cytokine levels also fluctuate significantly over a span of several months [17]. The fluctuation raises a problem regarding when to sample for a cytokine assay since the concentrations of a specific cytokine may vary largely between samples. This consideration led to our strategy to find cytokines with minimal fluctuation. On the other hand, we speculate that the fluctuation might reflect to some extent the change in the intensity of the immune response, and similar fluctuation patterns might suggest that relevant cytokines might be modulated similarly. For example, if cytokine A fluctuated with a pattern similar to that of cytokine B, this correlation might mean that A and B were regulated by the same stimulus or that A might be a stimulus or product of stimulation of B. Under this condition, picking A and B simultaneously as biomarkers would be at least partially redundant. This consideration led to our strategy to select one cytokine from one closely correlated group (Fig. 2).

The effectiveness of our strategy can be exemplified by the selection of M-CSF. This cytokine was clustered with IL-6 (Fig. 2B), indicating a relatively high correlation between them. Recent reports showed that IL-6 was associated with the severity of COVID-19 [10, 18]. According to our strategy, only 1 of the 2 cytokines would be desirable to minimize redundancy. Further analysis showed that M-CSF fluctuated less and classified different populations more accurately than IL-6 (Fig. 2), suggesting that M-CSF should be a better choice than IL-6.

Guided by the 2 strategies, we finally narrowed down the candidate cytokines to 3, M-CSF, IL-8 and SCF. A multiclass classification model constructed with these cytokines accurately predicted 77.4% of people in the healthy control group and asymptomatic and nonsevere groups for quasiindependent samples. However, this model was not successful in predicting severe cases, with an accuracy of only 1.7%, indicating the difficulty in finding cytokines that can discriminate the entire spectrum of COVID-19 simultaneously. We adjusted our strategy by screening for cytokines that showed significant differences between nonsevere and severe patients. This effort resulted in a single cytokine, HGF, which predicted severe patients with a significantly improved accuracy (84.6% vs 1.7%). This finding is consistent with 2 recent studies showing that the increase of HGF was associated with disease severity during COVID-19 progression [13, 14].

HGF is a potent antiinflammatory factor that ameliorates inflammation and dysfunction in a wide variety of experimental animal models [19]. It inhibits inflammation by interfering with proinflammatory NF-κB signaling [20, 21]. Considering its function, the increase in HGF in severe COVID-19 must not be a reason for but rather a consequence of excessive inflammation. Of note, HGF levels did not increase in asymptomatic patients and even decreased in nonsevere patients compared with those in healthy controls, suggesting that it might be unnecessary to upregulate HGF to control a moderate inflammatory response (Fig. 6). This result explains why HGF was ruled out from the candidate cytokines used to classify the 4 populations (Fig. 1). Presumably, HGF might significantly increase only when inflammation mounts toward an uncontrolled storm. If this is the case, HGF might be considered a curb of the cytokine storm and a candidate treatment for critical COVID-19 patients. Indeed, HGF treatment showed benefits for inflammatory bowel disease in an animal model and has been used to attenuate the inflammatory response to spinal cord injury in a clinical trial [22].

**Fig. 6.**
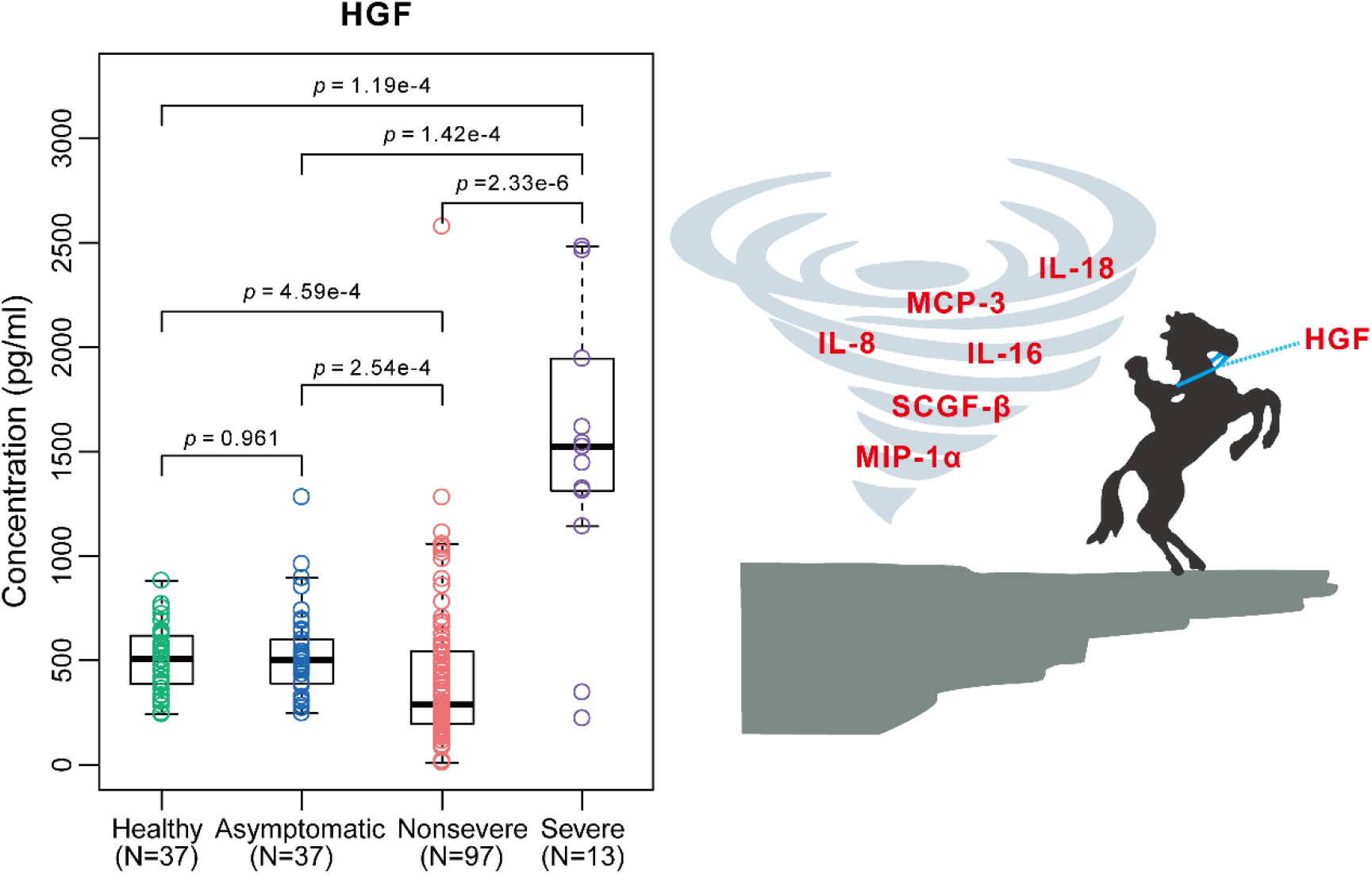
HGF might be a curb for cytokine storms in COVID-19 patients. HGF levels significantly decreased in the nonsevere patients but increased in the severe patients compared with those in the other 2 groups.

As suggested above, a covibration might imply a modulation relationship among cytokines: either the cytokines could be modulated by the same regulators or the cytokines could be regulators of the correlated cytokines. Interestingly, there was evidence supporting this hypothesis: an experiment conducted in canine endothelial cells demonstrated that M-CSF dose-dependently induced IP-10 mRNA expression, consistent with our finding that M-CSF fluctuated with a high correlation to IP-10 (r = 0.73, data not shown). This result implies the possibility that M-CSF could be a modulator of IP-10 in the immune response. Statistically high correlations also existed between M-CSF and IL-6 (r = 0.79) and IFN-γ and IL-2Rα (r = 0.73). It is interesting to investigate whether a functional relationship exists between these pairs of cytokines. A limitation of this study is that the sample size, especially that of severe patients, was small, and no critical and fatal cases were included. The accuracy of the cytokine biomarkers to classify populations with different states of SARS-CoV-2 infection needs to be tested further in a large and totally independent population. In addition, since all the cytokines were tested only when the patients had already been in corresponding states, it is not known how these cytokine biomarkers would predict which patients could develop severe illness.

## Data Availability

All the data are available from the corresponding author upon request.

## Main points

M-CSF, IL-8 and SCF together discriminated healthy individuals, asymptomatic infection subjects and nonsevere COVID patients;

HGF discriminated nonsevere and severe COVID-19, with an accuracy of 84.6% for a cutoff value of 1128 pg/ml;

Similar fluctuation patterns for M-CSF/IL-6/IP-10 and IFN-γ/IL-R2α might imply functional relationships among these cytokines; and HGF could be a candidate for critical COVID-19 therapy.

## Acknowledgments

This work was supported by the Emergency Project from the Science & Technology Commission of Chongqing; The Major National S&T program grant (2017ZX10202203 and 2017ZX10302201) from Science & Technology Commission of China.

---

**Table S1.** Demographics and baseline characteristics of the subjects in the cross-sectional analysis.

| Characteristics | Healthy<br>(N=37) | Asymptomatic<br>(N=37) | Mild (N=97) | Severe<br>(N=13) |
| --- | --- | --- | --- | --- |
| Age (median, IQR) | 35 (30-51) | 41 (27-50) | 46 (37-55) | 69 (54-79) |
| **Sex (Male/Female)** |  |  |  |  |
| Male | 16 (43.2%) | 15 (40.5%) | 52 (53.6%) | 6 (46.2%) |
| Female | 21 (56.8%) | 22 (59.5%) | 45 (46.4%) | 7 (53.8%) |
| **Exposure** |  |  |  |  |
| From wuhan | - | 9 (24.3%) | 40 (41.2%) | 7 (53.8%) |
| Close contacts | - | 28 (75.7%) | 57 (58.8%) | 6 (46.2%) |
| Days after exposure or<br>symptoms onset (median,<br>IQR) | - | 28.5 (28-30) | 9 (6-13) | 10 (9-19) |
| **Comorbidities** |  |  |  |  |
| Hypertension | - | 7 (13.5%) | 14 (14.4%) | 1 (7.7%) |
| Cardiovascular<br>disease | - | - | 3 (3.1%) | 1 (7.7%) |
| Diabetes | - | 1 (2.7%) | 6 (6.2%) | 2 (15.4%) |
| COPD | - | 0 (0.0%) | 1 (1.0%) | 1 (7.7%) |
| Chronic kidney<br>disease | - | 0 (0.0%) | 1 (1.0%) | 0 (0.0%) |
| Chronic liver disease | - | 2 (5.4%) | 3 (3.1%) | 1 (7.7%) |
| Tuberculosis | - | 0 (0.0%) | 2 (2.1%) | 0 (0.0%) |
| Any | - | 9 (24.3%) | 26 (26.8%) | 5 (38.5%) |
| **Signs and symptoms** |  |  |  |  |
| Fever | - | - | 47 (48.5%) | 6 (46.2%) |
| Fatigue | - | - | 18 (18.6%) | 7 (53.8%) |
| Dry cough | - | - | 44 (45.4%) | 6 (46.2%) |
| Anorexia | - | - | 9 (9.3%) | 7 (53.8%) |
| Myalgia | - | - | 12 (12.4%) | 2 (15.4%) |
| Dypnea | - | - | 5 (5.2%) | 6 (46.2%) |
| Expectoration | - | - | 22 (22.7%) | 5 (38.5%) |
| Pharyngalgia | - | - | 14 (14.4%) | 0 (0.0%) |
| Diarrhea | - | - | 7 (7.2%) | 2 (15.4%) |
| Nausea | - | - | 5 (5.2%) | 3 (23.1%) |
| Dizziness | - | - | 4 (4.1%) | 2 (15.4%) |
| Headache | - | - | 5 (5.2%) | 2 (15.4%) |
| Vomiting | - | - | 1 (1.0%) | 1 (7.7%) |
| Abdominal pain | - | - | 1 (1.0%) | 0 (0.0%) |
| Chill | - | - | 10 (10.3%) | 2 (15.4%) |
| Nasal congestion | - | - | 4 (4.1%) | 1 (7.7%) |
| Rhinorrhea | - | - | 8 (8.2%) | 0 (0.0%) |
| Chest stuffiness | - | - | 12 (12.4%) | 2 (15.4%) |

**Table S2.** Demographics and baseline characteristics of the subjects providing sequential samples.

| **Characteristics** | **Sequential serum patients**<br>**(N=38)** |
| --- | --- |
| Age (median, IQR) | 50 (44-63) |
| **Sex (Male/Female)** |  |
| Male | 20 (52.6%) |
| Female | 18 (47.4%) |
| **Exposure** |  |
| From wuhan | 8 (21.1%) |
| Close contacts | 30 (78.9%) |
| Days after exposure or symptoms onset (median, IQR) | 7 (5-10) |
| **Comorbidities** |  |
| Hypertension | 10 (26.3%) |
| Cardiovascular disease | 3 (7.9%) |
| Diabetes | 3 (7.9%) |
| COPD | 1 (2.6%) |
| Chronic kidney disease | 1 (2.6%) |
| Chronic liver disease | 2 (5.3%) |
| Tuberculosis | 1 (2.6%) |
| Any | 17 (44.7%) |
| **Signs and symptoms** |  |
| Fever | 17 (44.7%) |
| Fatigue | 6 (15.8%) |
| Dry cough | 12 (31.6%) |
| Anorexia | 3 (7.9%) |
| Myalgia | 6 (15.8%) |
| Dypnea | 4 (10.5%) |
| Expectoration | 7 (18.4%) |
| Pharyngalgia | 4 (10.5%) |
| Diarrhea | 2 (5.3%) |
| Nausea | 1 (2.6%) |
| Dizziness | 3 (7.9%) |
| Headache | 1 (2.6%) |
| Vomiting | 1 (2.6%) |
| Chill | 2 (5.3%) |
| Nasal congestion | 3 (7.9%) |
| Rhinorrhea | 3 (7.9%) |
| Chest stuffiness | 1 (2.6%) |

**Fig. S1.**
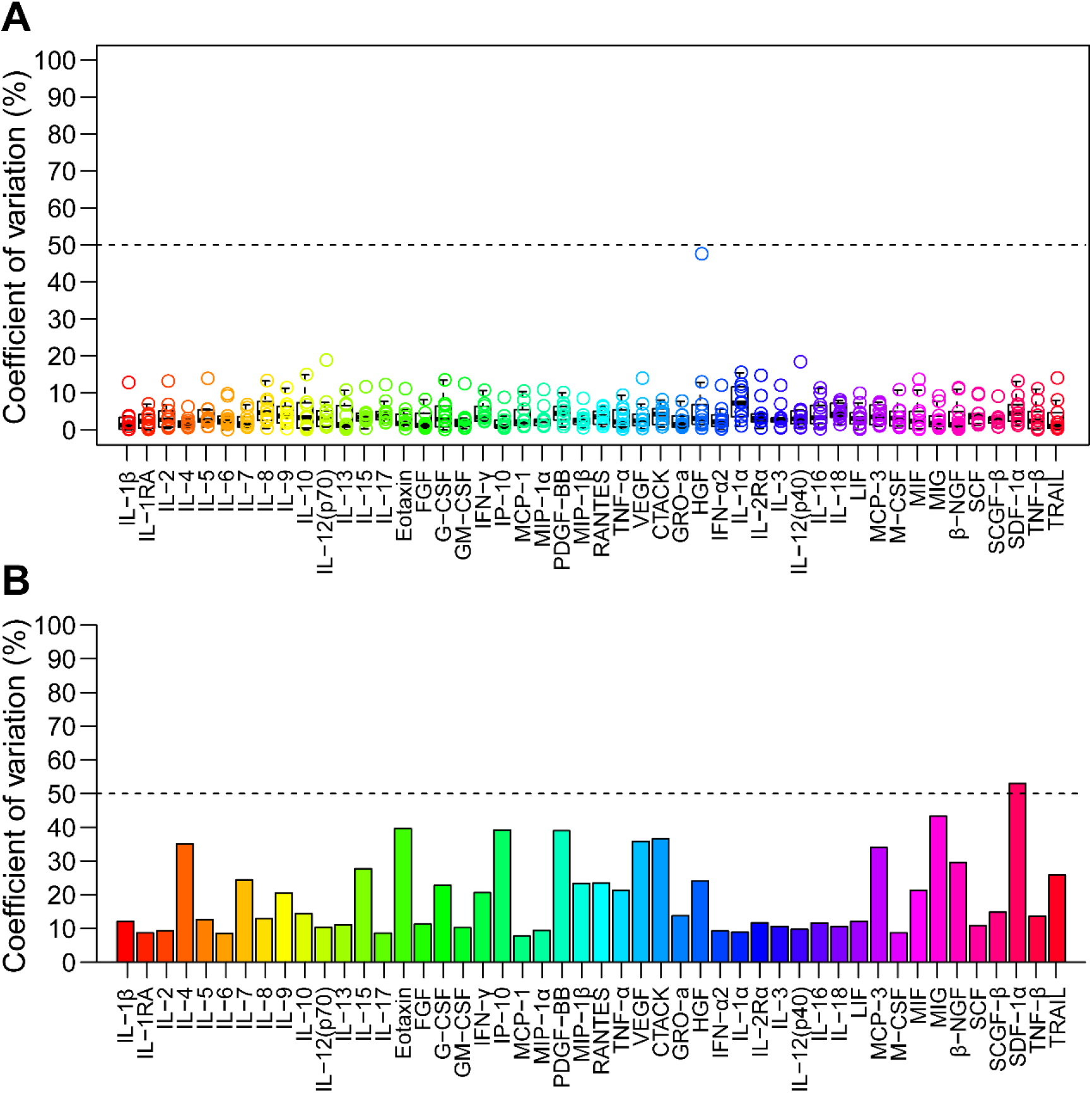
Coefficients of variation of cytokine controls in the assay. The CVs of the cytokine controls of each cytokine assayed in the same plate (A) or in different plates (B) are shown.

**Fig. S2.**
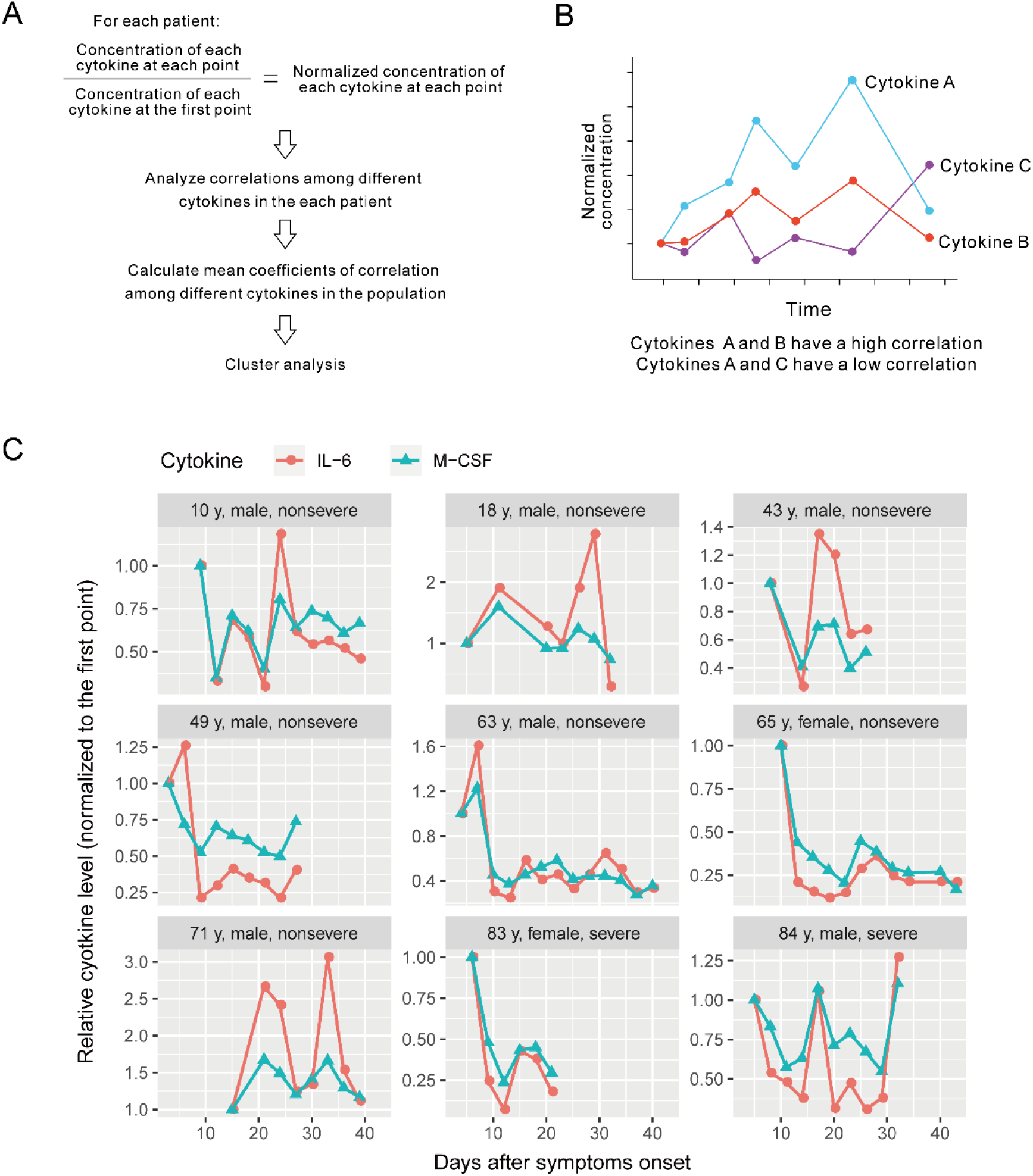
Some cytokines fluctuated with a similar pattern. The concentration of each cytokine from each patient was normalized against that of the first sample, and the fluctuation patterns were analyzed (A and B). C, Examples showing covibration in the fluctuations of IL-6 and M-CSF during illness. These examples also demonstrate generally higher amplitudes of fluctuation of IL-6 than those of M-CSF.

**Fig. S3.**
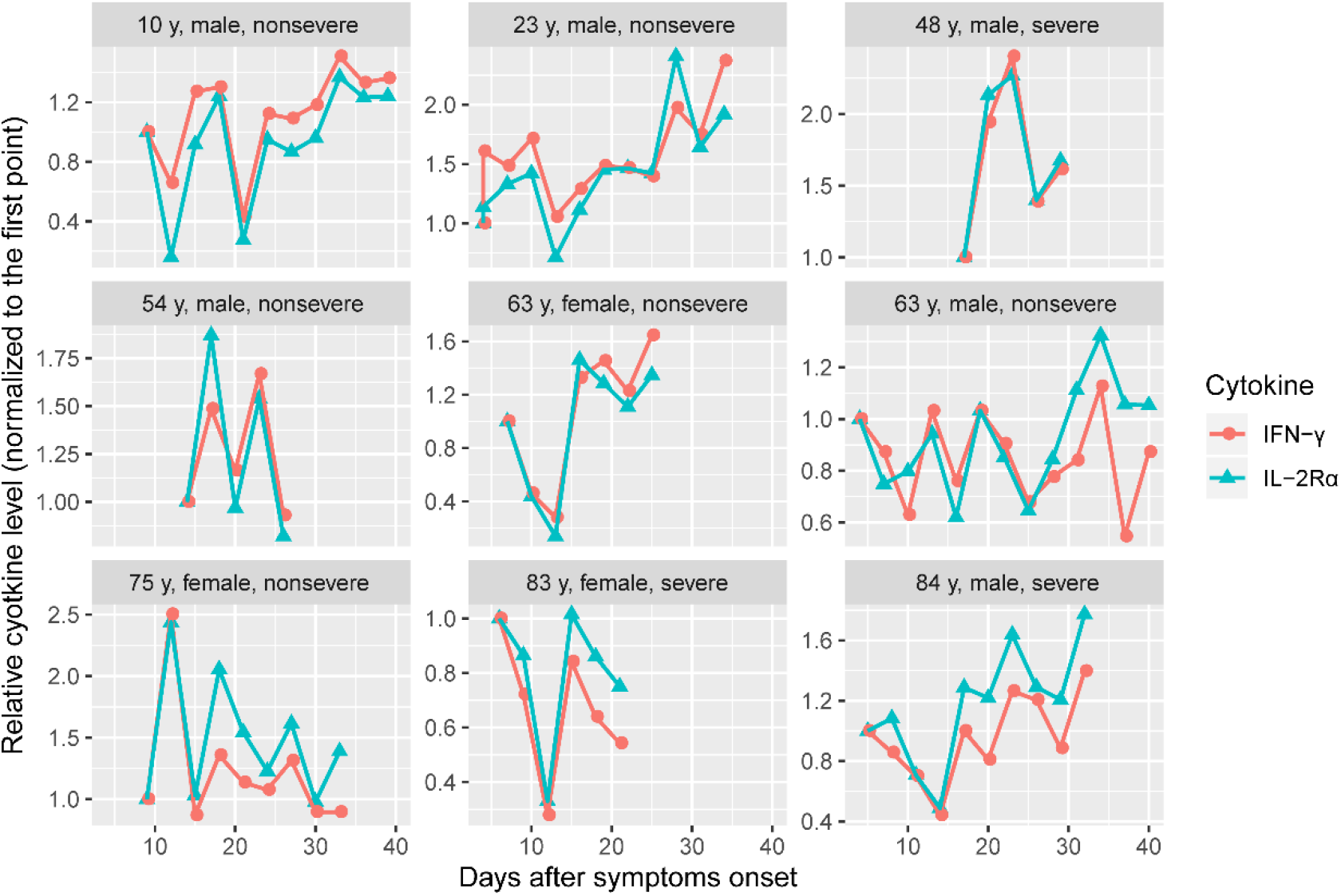
IFN-γ and IL-2Rα fluctuated with a similar pattern in some patients.

**Fig. S4.**
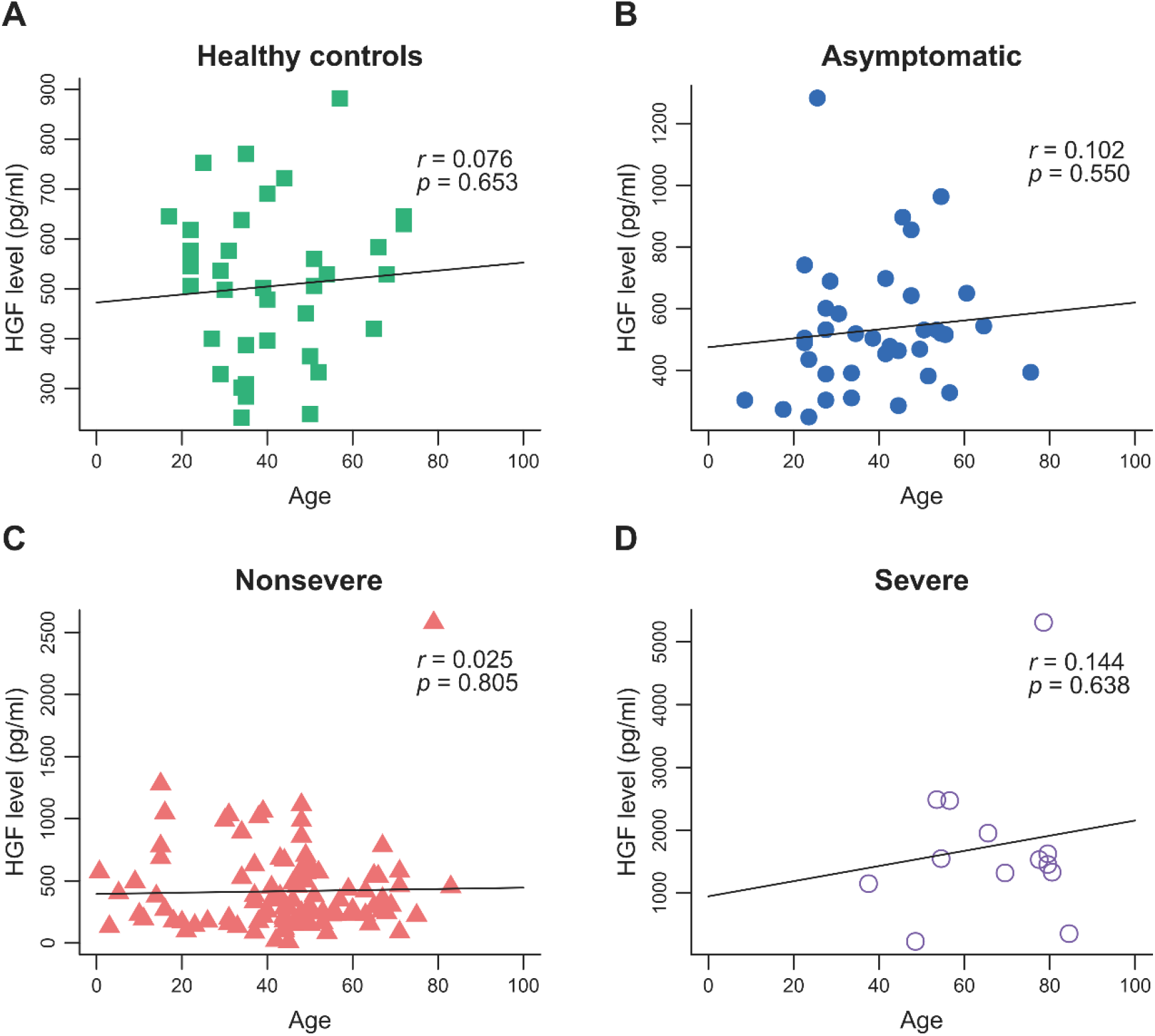
HGF levels are not associated with ages in different groups.

